# HLA-A*11:01:01:01, HLA*C*12:02:02:01-HLA-B*52:01:02:02, age and sex are associated with severity of Japanese COVID-19 with respiratory failure

**DOI:** 10.1101/2021.01.26.21250349

**Authors:** Seik-Soon Khor, Yosuke Omae, Nao Nishida, Masaya Sugiyama, Noriko Kinoshita, Tetsuya Suzuki, Michiyo Suzuki, Satoshi Suzuki, Shinyu Izumi, Masayuki Hojo, Norio Ohmagari, Masashi Mizokami, Katsushi Tokunaga

## Abstract

Severe acute respiratory syndrome coronavirus 2 (SARS-CoV-2), the virus causing coronavirus disease 2019 (COVID-19) was announced as an outbreak by the World Health Organization (WHO) in January 2020 and as a pandemic in March 2020. The majority of infected individuals have experienced no or only mild symptoms, ranging from fully asymptomatic cases to mild pneumonic disease. However, a minority of infected individuals develop severe respiratory symptoms. The objective of this study was to identify susceptible HLA alleles and clinical markers for the early identification of severe COVID-19 among hospitalized COVID-19 patients. A total of 137 patients with mild COVID-19 (mCOVID-19) and 53 patients with severe COVID-19 (sCOVID-19) were recruited from the Center Hospital of the National Center for Global Health and Medicine (NCGM), Tokyo, Japan for the period of February–August 2020. High-resolution sequencing-based typing for eight HLA genes was performed using next-generation sequencing. In the HLA association studies, HLA-A*11:01:01:01 [P_c_ = 0.013, OR = 2.26 (1.27–3.91)] and HLA-C*12:02:02:01-HLA-B*52:01:01:02 [P_c_ = 0.020, OR = 2.25 (1.24–3.92)] were found to be significantly associated with the severity of COVID-19. After multivariate analysis controlling for other confounding factors and comorbidities, HLA-A*11:01:01:01 [P = 3.34E-03, OR = 3.41 (1.50–7.73)], age at diagnosis [P = 1.29E-02, OR = 1.04 (1.01–1.07)] and sex at birth [P = 8.88E-03, OR = 2.92 (1.31–6.54)] remained significant. Early identification of potential sCOVID-19 could help clinicians prioritize medical utility and significantly decrease mortality from COVID-19.

## Introduction

Severe acute respiratory syndrome coronavirus 2 (SARS-CoV-2) is the virus responsible for the onset of COVID-19, and has so far infected more than 87 million people worldwide, with the global death toll surpassing 1.8 million (as of January 10, 2021). The majority of infected individuals have experienced only mild or minor symptoms, ranging from asymptomatic cases to mild pneumonic disease. However, a minority of infected individuals developed severe respiratory symptoms but eventually survive with the aid of mechanical ventilatory support.

As of January 10, 2021, mortality rates of COVID-19 (https://coronavirus.jhu.edu/data/mortality) were reported as highest in Mexico (8.8%), followed by Iran, Italy, Hungary, and Indonesia with approximate mortality rates of 3–4%, the United Kingdom, South Africa, Colombia, Canada, Spain, Brazil, France, Poland and Germany at around 2–2.9%, and other countries with reported mortality rates less than 2%. Differences in mortality rates can be caused by several factors, such as differences in the number of people tested, the capacity of local healthcare systems to handle increasing numbers of patients during the outbreak, the demographics of the population (with mortality rates known to be higher in older populations) and the different preventive measurements attempted by local governments. Nonetheless, while Japan stands as one of the oldest populations in the world (with citizens 65 years or older accounting for approximately 28% of the total population), the country has only suffered a mortality rate of 1.9%. Emerging clinical evidence has suggested that factors such as Bacillus Calmette-Guérin (BCG)-vaccination status [1], viral genomic strain [2, 3] and host genetic factors [4, 5] may be related to the severity and mortality of COVID-19.

The immunogenetic background of the host, such as HLA diversity, is well known to play an essential role in determining host responses to emerging infectious diseases such as TB [6], HBV [7–10], HCV [11, 12], HIV [13, 14], SARS-CoV-1 [15–19] and SARS-CoV-2 [20–22]. In cases of viral infection, viruses have successfully breached the early layers of the innate defense system (early nonspecific responses) such as fever, phagocytosis and inflammation. The human second line of defense is heavily reliant on the HLA-restricted T-cell response mechanism, in which viral epitopes are presented by dendritic cells to CD8+ T lymphocytes through interactions with HLA class I alleles and CD4+ T lymphocytes through HLA class II alleles. Viral epitope presentation by HLA class I leads to clonal expansion of HLA-restricted CD8+ cytotoxic T lymphocytes (CTLs), which are primed to perform antiviral defense during acute infection. Subsequently, latent reinfection and reactivation of the virus are controlled by memory CTLs.

The 2002–2003 SARS-CoV-1 outbreak is known to have higher pathogenicity and higher mortality rates, while SARS-CoV-2 infections show higher transmissibility. Experimental HLA association studies have indicated that HLA class I and class II alleles such as B*46:01 [15], B*07:03 [16], Cw*08:01 [17], and DRB1*12:02 [18] carriers are more susceptible to SARS-CoV-1 infection. Early studies of SARS-CoV-2 associations with HLA alleles generally used small sample sizes, but B*15:27 and C*07:29 have nonetheless been reported as associated with SARS-CoV-2 susceptibility in the Chinese population [20] and a SARS-CoV-2 susceptibility study with larger sample size in China reported associations of A*11:01, B*51:01 and C*11:01 with severity of COVID-19 in China [23]. On the other hand, B*27:07, DRB1*15:01, and DQB1*06:02 have been reported as susceptible HLA alleles in Italian COVID-19 [21].

Due to emerging evidence that the diversity of HLA allele distributions across different populations is related to the susceptibility to and severity of COVID-19, this study was designed to examine HLA alleles associated with the susceptibility and severity of Japanese COVID-19.

## Materials and Methods

### National Center for Global Health and Medicine (NCGM) COVID-19 patients

A total of 190 Japanese COVID-19 patients (all COVID-19 [aCOVID-19]) were recruited from the Center Hospital of the NCGM for the period of February–August 2020. In this study, severe COVID-19 (sCOVID-19) was defined according to the most recent (27 May 2020) clinical guidelines from the WHO (https://www.who.int/publications/i/item/clinical-management-of-covid-19), which defined “severe disease” as adults with clinical signs of pneumonia (fever, cough, dyspnea, fast breathing) accompanied by one of the following: respiratory rate > 30 breaths/min; severe respiratory distress; or peripheral oxygen saturation (SpO_2_) < 90% on room air. The sCOVID-19 patients comprised 27.9% of total patients who participated in this study. Written informed consent was obtained from all patients. This study was approved by the ethical committees from NCGM.

### Tokyo Healthy Control (THC)

THC samples comprised 423 healthy individuals residing in the Tokyo area before the COVID-19 outbreak. Informed consent was obtained from all of the THC sample participants. This study was approved by the ethical committees from NCGM.

### High-resolution HLA genotyping by next-generation sequencing

The AllType assay (One Lambda, West Hills, CA) was designed to cover the full length of five HLA genes (HLA-A, -C, -B, -DQA1 and -DPA1) and partial coverage for six HLA genes (HLA-DRB1, -DRB3, -DRB4, -DRB5, -DQB1 and -DPB1). Experimental protocols were carried out following vendor instructions, consisting of HLA target amplification, HLA library preparation, HLA template preparation, and HLA library loading onto an ion 530v1 chip (Thermo Fisher Scientific, Waltham, MA) in the ion chef (Thermo Fisher Scientific) followed by final sequencing on the ion S5 machine (Thermo Fisher Scientific).

### HLA genotype assignment

Demultiplexing of barcodes and base-calling was carried out in Torrent Suite version 5.8.0 (Thermo Fisher Scientific). Raw fastq reads were extracted using the FileExporter function in Torrent Suite version 5.8.0. HLA genotype assignments were carried out using two different types of software, namely HLATypeStream Visual (TSV v2.0; One Lambda, West Hills, CA) as the default software for the Alltype™ NGS Assay and NGSengine® (v2.18.0.17625) by the GenDX company (GenDX, Utrecht, the Netherlands). The default analysis parameters and healthy metrics threshold was applied for TSV2.0 while we applied the “ignore regions” function in the NGSengine to eliminate known sequencing error sites in the ion S5 system. For the fully covered HLA genes (HLA-A, -C, -B, -DQA1 and -DPA1), 4-field HLA alleles were determined after comparing the reads with the IMGT 3.40.0 database; 3-field HLA alleles were determined for partially covered HLA genes (HLA-DRB1, - DRB345, -DQB1 and -DPB1). Novel HLA alleles that are absent from the IMGT 3.40.0 database or ambiguous HLA alleles were subjected to Pacbio Sequel sequencing by H.U.Group Research Institute (Tokyo, Japan). After confirming the presence of novel HLA alleles using Pacbio Sequel sequencing, Pacbio consensus reads were submitted to GenBank and the IMGT nomenclature committee for the official naming of HLA alleles.

### Statistical analysis

Case-control HLA allele association tests, HLA haplotype estimations, case-control HLA haplotype association tests, Hardy-Weinberg equilibrium tests, and HLA amino acid association tests were prepared and analyzed using the Bridging ImmunoGenomics Data Analysis Workflow Gaps [24] (BIGDAWG) R package. The default parameters of BIGDAWG were used except for the manual specification of HLA haplotypes for testing. Logistic regression, univariate, and multivariate analyses of COVID-19 comorbidities were calculated using R statistics software v4.0.1 (R Foundation for Statistical Computing, Vienna, Austria).

To evaluate associations of risk HLA alleles and comorbidities with the severity of COVID-19, we applied univariate analysis (generalized linear model) to observe the association of associated risk HLA alleles, age, sex, and comorbidities such as high blood pressure, type 2 diabetes, obesity, respiratory diseases (chronic obstructive pulmonary disease [COPD], asthma, tuberculosis [TB]), hyperuricemia and dyslipidemia with the severity of COVID-19.

## Results

### Clinical characteristics of the 190 Japanese COVID-19 patients

Age distribution and other comorbidities in sCOVID-19 and mCOVID-19 patients are shown in Figure 1a and Figure 1b, respectively. Mean age at diagnosis for sCOVID-19 and mCOVID-19 were 57 years and 46 years, respectively, with males comprising 72% of sCOVID-19 patients and 46% of mCOVID-19 patients. In general, the most common comorbidities in sCOVID-19 were high blood pressure (43%, 23/53), type 2 diabetes (26%, 14/53), hyperuricemia (19%, 10/53), dyslipidemia (13%, 7/53), obesity (9%, 5/53) and COPD/asthma/TB (8%, 4/53). In comparison, comorbidities were shown to be lower in mCOVID-19 with high blood pressure (18%, 24/137), type 2 diabetes (8%, 11/137), hyperuricemia (4%, 5/137), dyslipidemia (12%, 17/137), obesity (5%, 7/137) and COPD/Asthma/TB (9%, 12/137).

**Figure 1:**
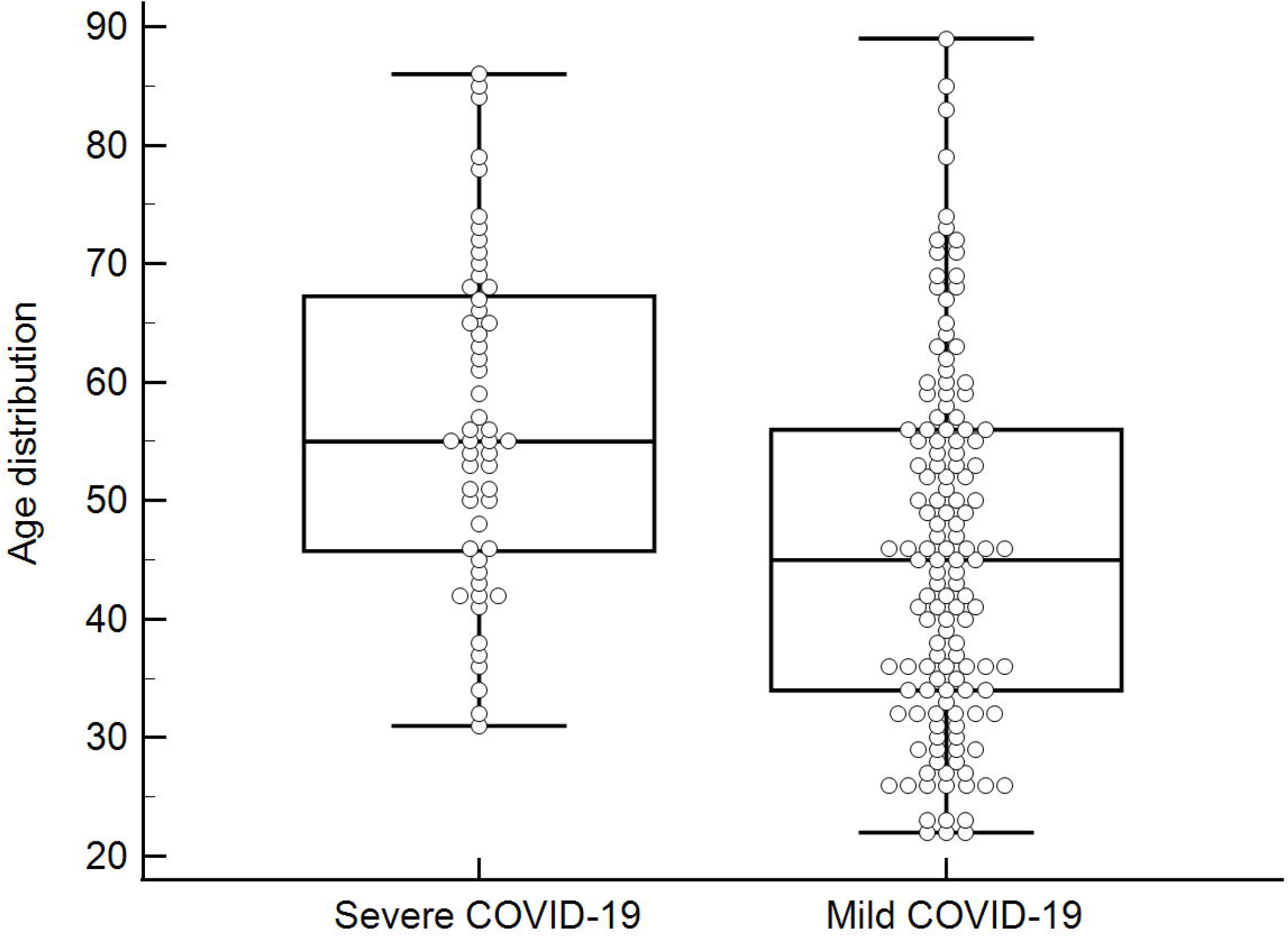
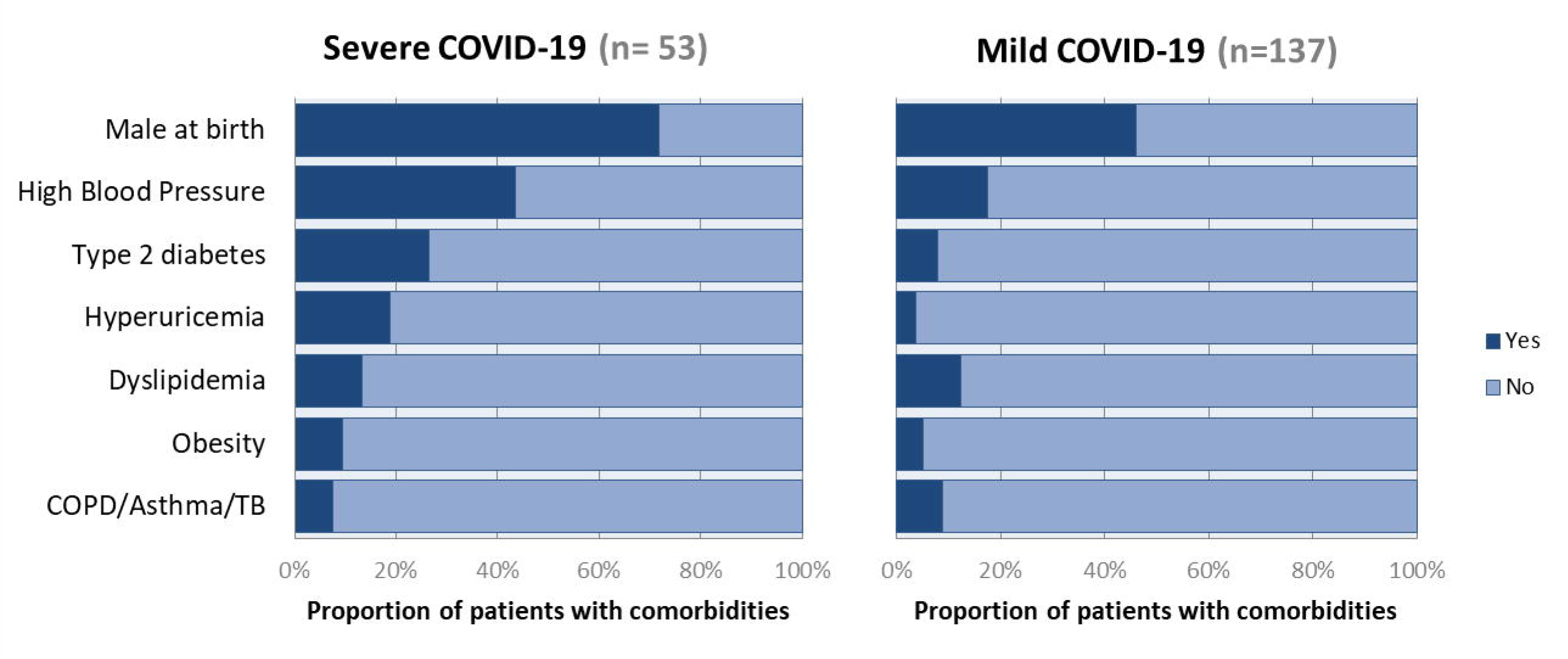
Distributions of age and existing comorbidities in sCOVID-19 and mCOVID-19. a) Age distribution of sCOVID-19 and mCOVID-19 b) Proportion of patients with comorbidities in sCOVID-19 and mCOVID-19

A comprehensive comparison of HLA allele associations for four different subgroups was performed in this study, namely: 1) aCOVID-19 versus healthy controls; 2) mCOVID-19 vs. healthy controls; 3) sCOVID-19 vs. healthy controls; and 4) sCOVID-19 vs. mCOVID-19.

### No association of HLA alleles with all COVID-19 (aCOVID-19 vs. controls)

For the identification of HLA alleles associated with SARS-CoV-2 infection, all 190 aCOVID-19 patients (comprising both mCOVID-19 and sCOVID-19) were compared with the 423 Japanese healthy controls. None of the HLA alleles in the 8 HLA genes tested showed significance after multiple corrections (Supplementary Table 1).

**Table 1:**
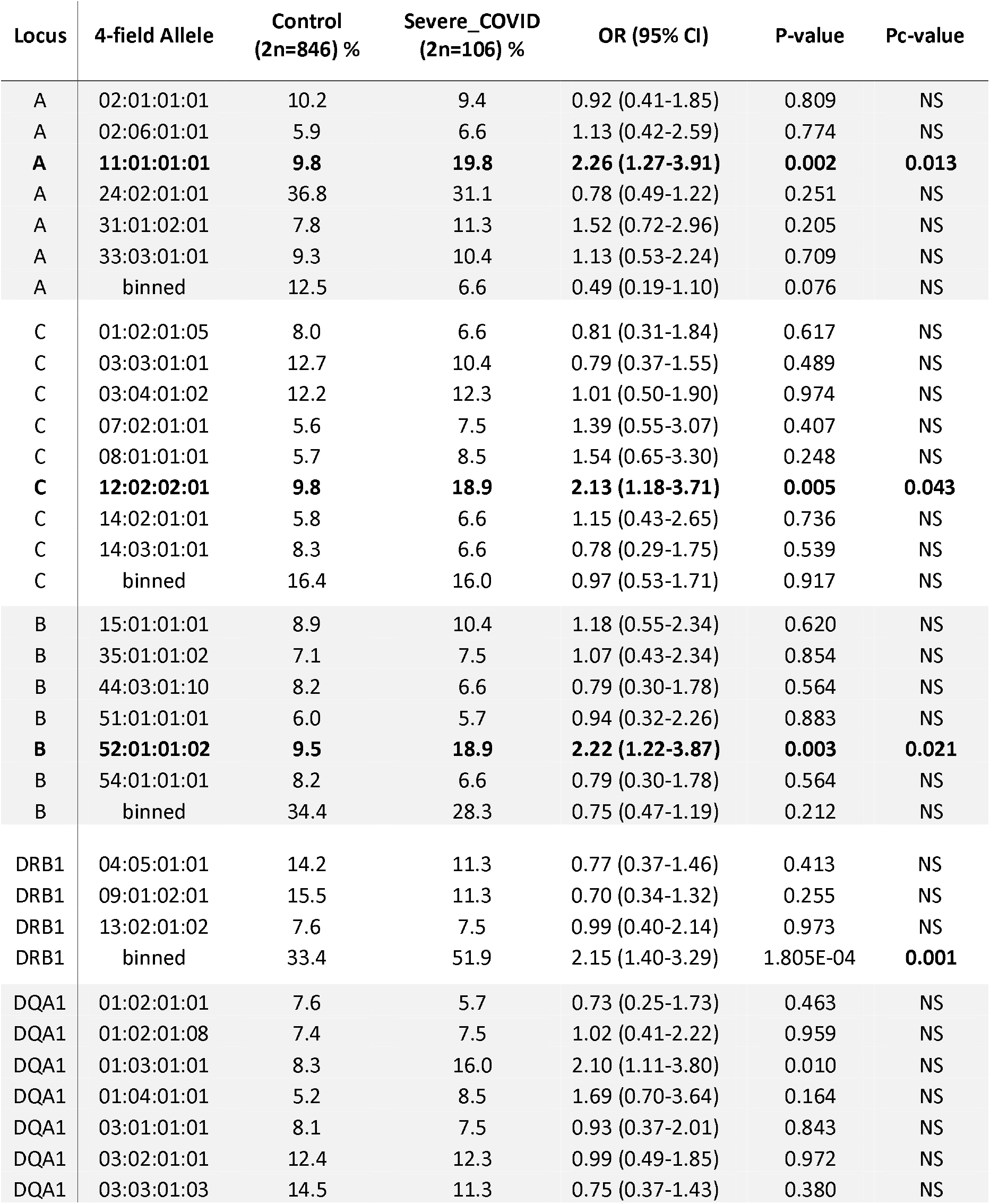
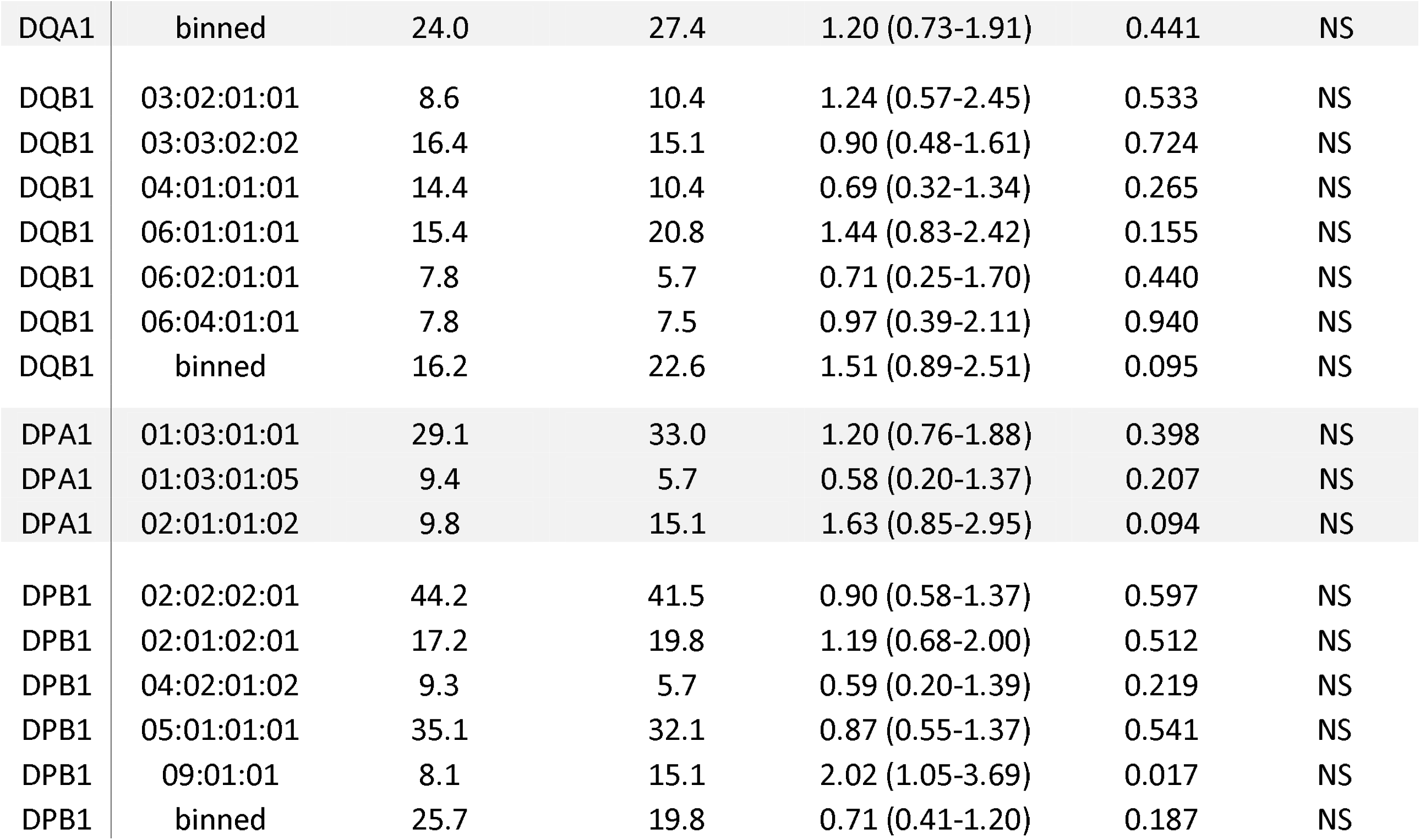
Association analysis of *HLA-A, -C, -B, -DRB1, -DQA1, -DQB1, -DPA1 and -DPB1* alleles in Japanese severe COVID-19 patients and Japanese healthy controls. Only HLA alleles with frequencies >5% are shown. Significant HLA alleles after multiple correction are highlighted in bold. Abbreviation:OR: Odd Ratio, 95% CI: 95% Confidence interval, Pc-value: multiple testing corrected pvalue, NS: Not significant binned: rare HLA alleles with expected count < 5 are combined into a common class

### No association of HLA alleles with mild COVID-19 (mCOVID-19 vs. controls)

To identify potential HLA alleles associated with mild symptoms of COVID-19, the 137 mCOVID-19 patients were compared with the healthy controls. No HLA alleles remained significant after multiple corrections (Supplementary Table 2). This result indicated that HLA frequencies for mCOVID-19 were similar to those of healthy controls.

**Table 2:** Comphrehensive HLA haplotype analysis of *HLA-A, -C* and *-B* in Japanese severe COVID-19 and Japanese healthy control. Significant HLA haplotypes after multiple correction are highlighted in bold. Abbreviation:OR: Odd Ratio, 95% CI: 95% Confidence interval, Pc-value: multiple testing corrected p-value, binned: rare HLA alleles with expected count < 5 are combined into a common class

| Locus | Allele | Control<br>(2n=846) % | Severe_COVID<br>(2n=106) % | OR (95% CI) | p.value | p.corrected |
| --- | --- | --- | --- | --- | --- | --- |
| A~C~B | 24:02:01:01~07:02:01:03~07:02:01:01 | 5.8 | 0.9 | 0.16 (0.00-0.93) | 0.035 | NS |
| **A~C~B** | **24:02:01:01~12:02:02:01~52:01:01:02** | **8.9** | **17.0** | **2.09 (1.12-3.72)** | **0.008** | **0.033** |
| A~C~B | 33:03:01:01~14:03:01:01~44:03:01:10 | 7.6 | 6.6 | 0.85 (0.32-1.93) | 0.703 | NS |
| A~C~B | binned | 77.7 | 75.5 | 0.89 (0.54-1.48) | 0.611 | NS |
| C~B | 01:02:01:05~54:01:01:01 | 7.9 | 6.6 | 0.83 (0.31-1.87) | 0.645 | NS |
| C~B | 03:03:01:01~35:01:01:02 | 5.9 | 4.7 | 0.79 (0.24-2.03) | 0.621 | NS |
| C~B | 03:04:01:02~40:02:01:01 | 5.2 | 3.8 | 0.71 (0.18-2.02) | 0.524 | NS |
| C~B | 07:02:01:03~07:02:01:01 | 6.8 | 3.8 | 0.54 (0.14-1.49) | 0.228 | NS |
| **C~B** | **12:02:02:01~52:01:01:02** | **9.4** | **18.9** | **2.25 (1.24-3.92)** | **0.003** | **0.020** |
| C~B | 14:02:01:01~51:01:01:01 | 5.8 | 5.7 | 0.98 (0.33-2.36) | 0.958 | NS |
| C~B | 14:03:01:01~44:03:01:10 | 8.2 | 6.6 | 0.79 (0.30-1.78) | 0.564 | NS |
| C~B | binned | 50.8 | 50.0 | 0.97 (0.63-1.48) | 0.875 | NS |
| A~C | 24:02:01:01~07:02:01:03 | 5.8 | 0.9 | 0.16 (0.00-0.93) | 0.035 | NS |
| **A~C** | **24:02:01:01~12:02:02:01** | **9.0** | **17.0** | **2.06 (1.11-3.67)** | **0.010** | **0.039** |
| A~C | 33:03:01:01~14:03:01:01 | 7.8 | 6.6 | 0.84 (0.32-1.90) | 0.674 | NS |
| A~C | binned | 77.4 | 75.5 | 0.90 (0.55-1.50) | 0.650 | NS |
| A~B | 24:02:01:01~07:02:01:01 | 5.9 | 0.9 | 0.15 (0.00-0.91) | 0.032 | NS |
| **A~B** | **24:02:01:01~52:01:01:02** | **9.0** | **17.0** | **2.06 (1.11-3.67)** | **0.010** | **0.039** |
| A~B | 33:03:01:01~44:03:01:10 | 7.6 | 6.6 | 0.85 (0.32-1.93) | 0.703 | NS |
| A~B | binned | 77.4 | 75.5 | 0.90 (0.55-1.50) | 0.650 | NS |

### Associations of HLA alleles with severe COVID-19 (sCOVID-19 vs. controls)

Severity of COVID-19 is directly related to the management of COVID-19 infection and also contributes to mortality from COVID-19. Association studies of the 8 HLA genes were carried out by comparing HLA allele frequencies in the 53 sCOVID-19 patients and 423 Japanese healthy controls (Table 1): HLA-A*11:01:01:01 [P_c_ = 0.013, OR = 2.26 (1.27–3.91)], HLA*C*12:02:02:01 [P_c_ = 0.043, OR = 2.13 (1.18–3.71)] and HLA-B*52:01:01:02 [P_c_ = 0.021, OR = 2.22 (1.22–3.87)] were significantly associated with the severity of COVID-19 after multiple corrections. To determine the independence of associated HLA alleles, HLA haplotype analyses were carried out. A total of 4 haplotypes remained significant after multiple corrections (Table 2), namely HLA-A*24:02:01:01-HLA-B*52:01:01:02 [P_c_ = 0.039, OR = 2.06 (1.11–3.67)], HLA-A*24:02:01-HLA-C*12:02:02:01 [P_c_ = 0.039, OR = 2.06 (1.11–3.67)], HLA-C*12:02:02:01-HLA-B*52:01:01:02 [P_c_ = 0.020, OR = 2.25 (1.24–3.92)] and HLA-A*24:02:01:01-HLA-C*12:02:01:01-HLA-B*52:01:01:02 [P_c_ = 0.033, OR = 2.09 (1.12–3.72)]. None of the HLA class II haplotypes or HLA class I - HLA class II haplotypes remained significant after multiple corrections (Supplementary Table 3). The most significantly associated HLA haplotype was HLA-C*12:02:02:01-HLA-B*52:01:01:02 [P_c_ = 0.020, OR = 2.25 (1.24–3.92)], and the level of significance decreased after including protective HLA-A*24:02:01:01 [P_c_ = 0.251, OR = 0.78 (0.49–1.22)] into the abovementioned haplotype, HLA-A*24:02:01:01-HLA-C*12:02:01:01-HLA-B*52:01:01:02 [P_c_ = 0.033, OR = 2.09 (1.12–3.72)]. Subsequent conditional analysis (Table 3) was performed on the statistically significant HLA alleles, HLA-A*11:01:01:01, HLA*C*12:02:02:01 and HLA-B*52:01:01:02. Each of HLA-A*11:01:01:01 and HLA*C*12:02:02:01 and HLA-B*52:01:01:02 were found to be independently associated with severity of COVID-19. All HLA*C*12:02:02:01 alleles were shown to be tightly linked with HLA-B*52:01:01:02 alleles and only a small proportion of A*11:01:01:01 alleles were linked with HLA*C*12:02:02:01 and HLA-B*52:01:01:02 haplotypes (Figure 2).

**Table 3:**
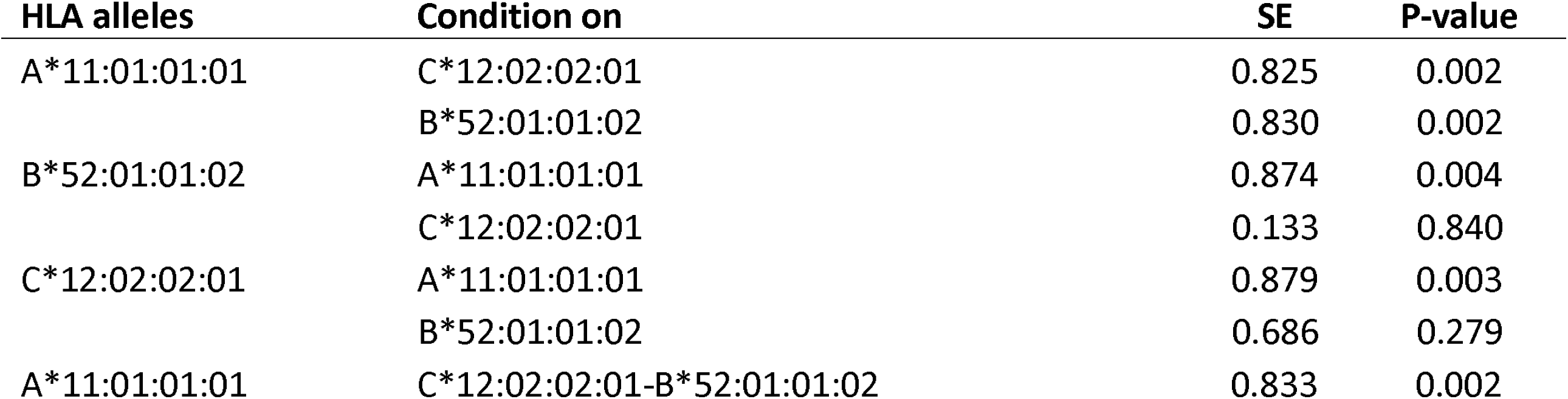
Conditional analysis on top associated HLA alleles and HLA haplotypes with the severity of COVID-19. Abbreviation:SE, Standard error.

**Figure 2:**
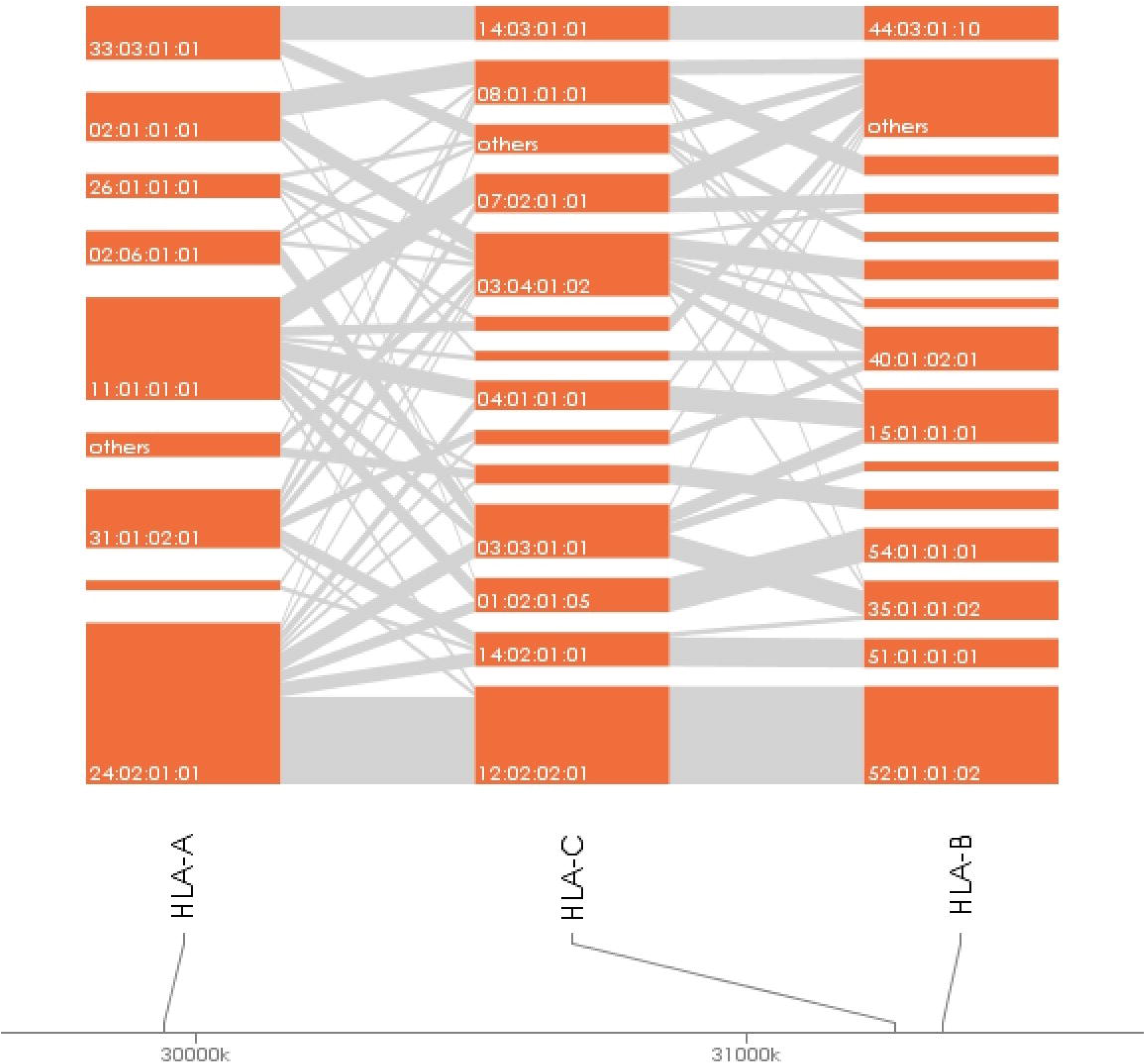
HLA allele linkage distributions of HLA-A, HLA-B and HLA-C in sCOVID-19.

### Associations of HLA alleles with severe COVID-19 (sCOVID-19 vs. mCOVID-19)

To identify potential HLA markers that can be used to prioritize treatment for early hospitalized COVID-19 patients, we compared HLA allele frequencies between the 53 sCOVID-19 patients and the 137 mCOVID-19 patients (Supplementary Table 4).

**Table 4:** Univariate analysis (Generalized Linear Model) of associated HLA alleles and comorbidities with the severity of COVID-19.

| **Factors** | **Univariate**<br>**p-value** | **OR (95% CI)** | **Multivariate**<br>**p-value** | **OR (95% CI)** |
| --- | --- | --- | --- | --- |
| A*11:01:01:01 | **1.71E-03** | 3.09 (1.52-6.25) | **3.34E-03** | 3.41 (1.50-7.73) |
| C*12:02:02:01-B*52:01:01:02 | 7.42E-02 | 1.75 (0.95-3.22) | - | - |
| Age | **3.52E-05** | 1.05 (1.03-1.07) | **1.29E-02** | 1.04 (1.01-1.07) |
| Sex | **1.83E-03** | 2.96 (1.50-5.91) | **8.88E-03** | 2.92 (1.31-6.54) |
| High_Blood_Pressure | **3.22E-04** | 3.61 (1.79-7.26) | 5.25E-01 | 1.33 (0.55-3.25) |
| Type2Diabetes | **1.40E-03** | 4.11 (1.73-9.79) | 1.00E-01 | 2.30 (0.85-6.18) |
| Obesity | 2.79E-01 | 1.93 (0.56-6.39) | - | - |
| COPD_Asthma_TB | 7.88E-01 | 0.85 (0.26-2.76) | - | - |
| Hyperuricemia | **1.60E-03** | 6.14 (1.99-18.96) | 7.93E-02 | 3.04 (0.88-10.50) |
| Dyslipidemia | 8.82E-01 | 1.07 (0.42-2.76) | - | - |

HLA*C*12:02:02:01 [P_c_ = 0.022, OR = 2.57 (1.29–5.09)] carrier was significantly associated with increased severity of COVID-19 patients, while due to the smaller effect size, HLA*C*12:02:02:01 and HLA-B*52:01:01:02 showed lower significance in comparison to the 53 sCOVID-19 patients and 423 Japanese healthy controls (Table 1).

## Discussion

We conducted a high-resolution sequencing-based typing of eight HLA genes to evaluate the association of HLA alleles/haplotypes with SARS-CoV-2 infection susceptibility and the severity of COVID-19. A total of 53 sCOVID-19 patients and 137 mCOVID-19 patients were recruited from the NCGM, Tokyo, Japan, from February to August 2020 and 423 previously recruited healthy controls were used as healthy comparators in this study.

To evaluate HLA alleles potentially associated with susceptibility to SARS-CoV-2 infection, aCOVID-19 (Supplementary Table 1) and mCOVID-19 (Supplementary Table 2) were compared with healthy controls, respectively. We found that no HLA alleles remained significant after Bonferroni correction, suggesting that no HLA alleles were either protective or conferred susceptibility to SARS-CoV-2 infection.

Patients with sCOVID-19 are tightly associated with COVID-19 mortality. We observed that the HLA-A*11:01:01:01 [P_c_ = 0.013, OR = 2.26 (1.27–3.91)] allele and HLA-C*12:02:02:01-HLA-B*52:01:01:02 [P_c_ = 0.020, OR = 2.25 (1.24–3.92)] haplotype were significantly associated with the severity of COVID-19. A recent publication by Wang et al. [23] reported significant associations of HLA-A*11:01, HLA-B*51:01, and HLA-C*14:02 with severe or critically severe Chinese COVID-19 patients compared to mild or moderate Chinese COVID-19 patients. Accordingly, HLA-A*11:01 was significantly associated with both Japanese and Chinese sCOVID-19 patients, while HLA-C*12:02:02:01-HLA-B*52:01:01:02 and HLA-B*51:01-HLA-C*14:02 were associated in Japanese and Chinese, respectively. The HLA associations of HLA-A*11:01 and HLA-C*12:02-HLA-B*52:01 are of particular importance, as Asian populations carry relatively high frequencies of these HLA alleles. Referring to the Allele Frequency Net Database (AFND) 2020 [25], HLA-C*12:02-HLA-B*52:01 is commonly present in Japanese with a frequency of 10.5%, followed by Asian populations residing in the United States (1.5–3.4%). On the other hand, HLA-A*11:01 is predominant in South-east Asia (17.7–61.3%), Japan (8.2–11.1%), China (16.2– 61.3%) and the Oceania region (2.6–63.6%). We expect that HLA-A*11:01 and HLA-C*12:02-HLA-B*52:01 could potentially act as a predictive marker for the severity of COVID-19 in the Asia region.

However, we could not replicate results for B*46:01, which has been predicted to have the fewest predicted binding peptides for SARS-CoV-2, thus Individuals carrying B*46:01 are predicted to be more vulnerable to SARS-CoV-2 infection [26]. Further, we could not confirm the findings of an HLA-binding prediction study [3] that predicted HLA-A*11:01, HLA-A*02:06, and HLA-B*54:01 as protective against SARS-CoV-2.

All samples used in this study were collected in the Center Hospital of the NCGM for the period from February to August 2020, where the mutant strain of SARS-CoV-2 was not yet prevalent in Japan. However, further studies are needed to evaluate correlations between the emerging mutant strain of SARS-CoV-2 and severity of COVID-19.

## Data Availability

The data collected in this study shall be available upon request

## Acknowledgements

We express our gratitude to Ms. Yoshimi Shigemori and Ms. Ayumi Nakayama for their technical assistance. We thank Dr.Ito Ikue and Dr.Sato Tetsuya from H.U.Group Research Institute for their technical support for Pacbio Sequel sequencing. This research is supported by Japan Agency for Medical Research and Development (AMED) under Grant Number JP20kk0205012 and JP20fk0108104 and the NCGM Intramural Research Fund 20A2002D.

## Notes

### Competing Interest Statement

The authors have declared no competing interest.

### Author Declarations

This study was approved by the ethical committees from National Center for Global Health and Medicine.

